# Physiological subphenotypes of ARDS: Prognostic and predictive enrichment for PEEP strategy

**DOI:** 10.64898/2026.03.27.26349397

**Authors:** G. Meza-Fuentes, I. Delgado, M. Barbé, I. Sánchez-Barraza, D. Filippini, M. Smit, J. Sinnige, L. Kramer, J. Smit, A Jonkman, M. Meade, M. A. Retamal, R. López, L.D.J. Bos

**Affiliations:** Instituto de Ciencias e Innovación en Medicina, Universidad del Desarrollo - UDD, Las Condes, Chile; Centro de Epidemiologia y Políticas de Salud, Universidad del Desarrollo - UDD, Las Condes, Chile; Intensive Care, Amsterdam UMC, University of Amsterdam, Amsterdam, Netherlands; Intensive Care, Erasmus MC, Rotterdam, Netherlands; Intensive care, McMaster University, Canada; Programa de Comunicación Celular en Cáncer, Instituto de Ciencias e Innovación en Medicina (ICIM). Clínica Alemana Universidad del Desarrollo - UDD - Santiago, Chile; Departamento de Paciente Crítico, Clínica Alemana, Vitacura, Chile; Grupo Intensivo, Instituto de Ciencias e Innovación en Medicina (ICIM), Facultad de Medicina, Clínica Alemana, Universidad del Desarrollo, Santiago, Chile

## Abstract

**Background:** Acute respiratory distress syndrome (ARDS) is characterised by substantial physiological heterogeneity, which contribute to a very variable clinical outcomes and therefore inconsistent responses to ventilatory strategies. We aimed to externally validate physiological ARDS subphenotypes previously identified using routine ventilatory and gas-exchange variables, assess their prognostic relevance across independent cohorts, and examine heterogeneity of treatment effect according to PEEP strategy.

**Methods:** Unsupervised Gaussian Mixture Modelling was used to identify physiological subphenotypes based on ventilatory mechanics and gas-exchange parameters. Labels were subsequently used to train and validate supervised classifiers using XGBoost. Prognostic relevance was assessed across three independent cohorts, including two randomised controlled trials (ALVEOLI and LOVS). Predictive enrichment for PEEP strategy was evaluated using individual patient data from ALVEOLI and LOVS (n = 1,532) using intention-to-treat analyses, applying both one-stage and two-stage fixed-effects IPD meta-analytic approaches to test for interaction between physiological subphenotype and PEEP strategy.

**Results:** Two distinct physiological subphenotypes, termed Efficient and Restrictive, were replicated across independent cohorts. Across each cohort, patients classified as Restrictive consistently exhibited higher all-cause 28-day mortality compared to Efficient patients. When pooled across studies, the Restrictive subphenotype was associated with a significantly increased risk of death (pooled odds ratio 1.75, 95% CI 1.36–2.24), with no evidence of between-study heterogeneity. Predictive analyses showed a statistically significant interaction between physiological subphenotype and PEEP strategy in the one-stage IPD model (p for interaction = 0.037), with concordant findings in the two-stage fixed-effects IPD meta-analysis (interaction OR 1.91, 95% CI 1.00–3.66; I² = 0%). Higher PEEP was associated with increased mortality in Efficient patients and reduced mortality in Restrictive patients, indicating effect modification by physiological subphenotype.

**Interpretation:** Physiological ARDS subphenotypes derived from routinely collected bedside data provide robust and externally validated prognostic stratification across observational and randomised trial cohorts. The observed interaction with PEEP strategy suggests that underlying physiological profiles may influence treatment response, supporting the concept that physiology-based be a starting point for personalized medicine and therefore better ventilatory strategies in future clinical trials.

## Introduction

Acute Respiratory Distress Syndrome (ARDS) is a common cause of respiratory failure in the intensive care unit (ICU) with a high mortality (1,2). In unselected patients, randomized clinical trials have largely failed to demonstrate consistent therapeutic benefit of well-developed interventions, a limitation increasingly attributed to inadequate patient stratification (3). ARDS can result in reduced respiratory system compliance, increased dead space ventilation, and impaired oxygenation, yet there are broadly varying patterns of physiological abnormalities (4–7). Despite this marked physiological diversity, ARDS patients continue to be treated with largely uniform interventions (8,9). The European Society of Intensive Care Medicine (ESICM) has identified heterogeneity as a central obstacle to therapeutic progress in ARDS, emphasizing that treatment effects may be diluted in broadly defined populations (10). There is now a broad consensus that we need better stratification approaches to align physiological patterns with appropriate treatments (3,4).

Individual physiological variables have long been used to define disease severity and stratify risk in ARDS. For example, respiratory system compliance (Crs) has been shown to moderate the effect of low tidal volume ventilation and a reduction in driving pressure (DP) seems to mediate the potential benefit of a higher positive end-expiratory pressure (PEEP) strategy (11). However, reliance on single physiological metrics is inherently limited due to the complex interactions between these parameters, whereby treatment decisions may significantly alter one variable without changing the risk for ventilator-induced lung injury (VILI). Instead, multidimensional physiological approaches that integrate respiratory mechanics, gas exchange, and ventilatory demand have been adopted as a more accurate way to characterize disease severity and the underlying pathophysiological complexity of ARDS (5,12).

Learning from the advances in the identification and validation of data-driven approaches to identify biological subphenotypes of ARDS, we used a comprehensive set of physiological variables to identify two physiological subphenotypes. The *Restrictive phenotype* exhibited marked impairment of respiratory mechanics, higher dead space, poorer oxygenation, and a substantially higher mortality risk (13), while the *Efficient subphenotype* maintained relatively normal physiology. These were truly latent classes as no single variable or simple combination of values could explain the allocation to one subphenotype, representing the complex interaction between decreased compliance, ineffective ventilation and limitation of tidal volume and driving pressure.

Despite a strong mechanistic rationale supporting its role in preventing alveolar collapse and reducing VILI (14,15), randomized trials comparing higher versus lower PEEP strategies have consistently failed to demonstrate a uniform survival benefit across unselected ARDS cohorts (16–18). Patient selection on single variables has also failed to identify consistent reductions in mortality (19). Mechanical ventilation strategies targeted at the underlying lung morphology showed potential benefits, but challenges in accurate classification led to treatment misalignment and possible harm (20). These observations suggest that the neutral results of PEEP trials do not reflect an absence of physiological effect, but rather the averaging of divergent responses across physiologically distinct patients, reinforcing the need for subphenotype-guided approaches to ventilatory management in ARDS.

Against this background, we investigated whether the previously described restrictive and efficient physiological subphenotypes capture stable, reproducible patterns of lung dysfunction that translate into differential responses to ventilatory management. To address this, we aimed to (1) externally validate the physiological and prognostic differences between the phenotypes, and (2) evaluate if there is heterogeneity of treatment effects (HTE) for low or high PEEP strategies between the phenotypes, using data from randomized controlled trials. We hypothesized that patients with a restrictive phenotype benefit from a higher PEEP strategy, while patients with an efficient phenotype do not.

## Methods

### 1.1 Study Populations and Cohorts

This study used individual patient data from four cohorts, for derivation, validation and evaluation of HTE. All studies included patients with ARDS according to the Berlin or American-European Consensus Conference (AECC) criteria, undergoing invasive mechanical ventilation in a mandatory mode. Patients after lung transplantation, with chronic respiratory disease such as obstructive diseases or fibrosis and on VV-ECMO were excluded. Derivation of the phenotypes was performed on 224 patients from a Chilean ICU registry. External validation was performed in 494 patients from Amsterdam UMC. To evaluate prognostic performance and HTE for PEEP, a parsimonious classifier model for physiological phenotypes model was applied to individual patient data from two landmark randomized controlled trials, Assessment of Low tidal Volume and elevated End-expiratory volume to Obviate Lung Injury (ALVEOLI, n= 549) and Lung Open Ventilation to Decrease Mortality in the Acute Respiratory Distress Syndrome (LOVS, n= 983).

In both ALVEOLI and LOVS, patients were randomized to protocolized lower versus higher PEEP strategies defined a priori by trial-specific ventilatory protocols (16,17). In ALVEOLI, both treatment groups received lung-protective ventilation with target tidal volumes of 6 mL/kg of predicted body weight and plateau airway pressures limited to ≤30 cm H₂O. The lower PEEP group was managed using a PEEP–FiO₂ table specifying relatively lower levels of PEEP for a given FiO₂, whereas the higher PEEP group followed a protocolized PEEP–FiO₂ table applying systematically higher PEEP levels across FiO₂ categories, without the routine use of recruitment maneuvers.

In the LOVS trial, the control strategy similarly targeted tidal volumes of 6 mL/kg of predicted body weight and plateau airway pressures ≤30 cm H₂O, using conventional levels of PEEP. In contrast, the experimental strategy combined higher levels of PEEP with the use of recruitment maneuvers and allowed plateau pressures up to 40 cm H₂O, reflecting a more aggressive lung recruitment approach. Thus, in LOVS, the higher PEEP strategy differed not only by higher end-expiratory pressures but also by the inclusion of recruitment maneuvers and higher allowable inspiratory pressures.

Although the specific ventilatory protocols differed between ALVEOLI and LOVS, both trials contrasted two clearly distinct strategies characterized by lower versus higher applied PEEP throughout mechanical ventilation. Treatment assignment was determined by randomisation within each trial and was independent of physiological subphenotype classification.

### 1.2 Phenotype derivation

#### 1.2.1 Input variables

In the original Chilean derivation cohort, appropriate variables for machine learning models were selected by eliminating redundant and collinear variables. Pearson correlation coefficients were used to identify highly collinear pairs (correlation coefficient > 0.8), and one variable from each pair was removed based on clinical interpretability. To further address redundancy, variables were evaluated using R² values to quantify their predictability by other variables, and those contributing minimal unique information were excluded.

The final set of ventilatory and gas-exchange variables included PEEP, peak pressure (Ppeak), plateau pressure (PPlat), respiratory rate (RR), end-tidal carbon dioxide (EtCO₂), carbon dioxide production (VCO₂), arterial carbon dioxide tension (PaCO₂), arterial oxygen tension (PaO₂), PaO₂/FiO₂ ratio, driving pressure (DP), tidal volume normalised to predicted body weight (Vt/PBW), normalised mechanical power (nMP) (21), respiratory system compliance (Crs), and the physiological dead space fraction (VD/VT), calculated using the Enghoff modification of the Bohr equation (22). All variables were obtained within the first 24 hours of invasive mechanical ventilation.

#### 1.2.2 Mixture model

Following variable selection, dimensionality reduction was performed using principal component analysis to address multicollinearity and enhance cluster separability. The resulting components were used as input for an unsupervised Gaussian Mixture Model (GMM). Model selection criteria, including the elbow method and silhouette analysis, supported a two-cluster solution, defining the Efficient and Restrictive physiological subphenotypes.

In the present study, the pretrained GMM was applied using the *predict* function to assign patients from external cohorts to subphenotypes based on posterior class probabilities.

### 1.3 External validation

External validation assessed the reproducibility of the previously derived unsupervised physiological subphenotypes. The original GMM, trained in the Chilean derivation cohort, was applied to the Amsterdam UMC dataset using an identical preprocessing pipeline, including variable definitions, scaling, normalisation, and plausibility filtering. A schematic overview of the model development, external validation, and application to treatment response is provided in Supplementary Figure S1.

### 1.4 Parsimonious classifier

To enable application of the subphenotyping framework in randomised trial datasets, a supervised classification model was subsequently developed. GMM-derived subphenotype labels were used as reference classes to train an Extreme Gradient Boosting (XGBoost) classifier based on routinely available ventilatory and gas-exchange variables. To ensure applicability in cohorts without capnography, ventilatory ratio (VR) (23) was used as a surrogate of physiological dead space (VD/VT). Variable selection was performed using recursive feature elimination with cross-validation (RFECV) to obtain a parsimonious and stable model (Supplementary Figure S2). Details of model training and hyperparameter selection are provided in Supplementary Table S1.

Classification performance was evaluated in the derivation cohort by comparing supervised predictions against the original GMM-derived subphenotype labels. Model performance was assessed using discrimination and classification metrics, including the area under the receiver operating characteristic curve (AUROCC), accuracy, and F1 score. Model training used a binary logistic objective function (log loss), with hyperparameters selected to balance model complexity and generalisability.

### 1.5 Outcomes

The primary outcome was all-cause mortality at day 28. Secondary analyses described differences in ventilatory and gas-exchange variables across physiological subphenotypes and assessed prognostic and predictive enrichment using 28-day mortality as the clinical endpoint.

### 1.6 Heterogeneity of treatment effect analysis

HTE was evaluated using individual patient data from the ALVEOLI and LOVS randomised trials. Patients were assigned to physiological subphenotypes using the supervised classification model based on pre randomisation baseline variables.

As an initial descriptive step, crude 28-day mortality was compared across subphenotypes and PEEP strategies in the pooled trial population. Subsequently, a one-stage IPD analysis was performed using a logistic regression model fitted to the combined dataset, including treatment group, physiological subphenotype, their interaction term, and adjustment for study membership. The interaction between treatment assignment and subphenotype was used to formally assess HTE.

To further assess consistency of treatment effects across trials, study-specific logistic regression models were fitted separately within each trial to estimate the effect of higher versus lower PEEP within each subphenotype. These study-specific estimates were subsequently pooled using a fixed-effects two-stage IPD meta-analysis to obtain pooled treatment effects within subphenotypes and a meta-analytic interaction estimate, defined as the difference between the log odds ratios for higher versus lower PEEP in the Efficient and Restrictive subphenotypes, in accordance with methodological recommendations for IPD interaction analyses (24,25).

All HTE analyses were pre-specified and focused on physiological subphenotype as the sole effect modifier.

### 1.7 Statistical analysis

Continuous variables were summarized as median and interquartile range (IQR), and categorical variables as counts and percentages. Baseline and physiological characteristics were compared between subphenotypes using the Mann–Whitney U test for continuous variables and the χ² test or Fisher’s exact test for categorical variables, as appropriate.

For supervised classification and HTE analyses, model performance metrics and analytical approaches were applied as detailed in the sections above. Treatment effects are reported as odds ratios with 95% confidence intervals, and between-study heterogeneity was assessed using the I² statistic. Marginal predicted probabilities were derived from fitted logistic regression models using estimated marginal means.

All statistical tests were two-sided, and a p value < 0.05 was considered statistically significant. Analyses were performed using Python (version 3.10.12) and R (version 4.4.2).

## Results

### 1.8 External validation

#### 1.8.1 Baseline and physiological characteristics

In the Amsterdam UMC cohort (n = 494), 153 patients (31%) were classified as Efficient and 341 (69%) as Restrictive. Baseline demographic characteristics were broadly similar between groups (Table 1).

**Table 1.**
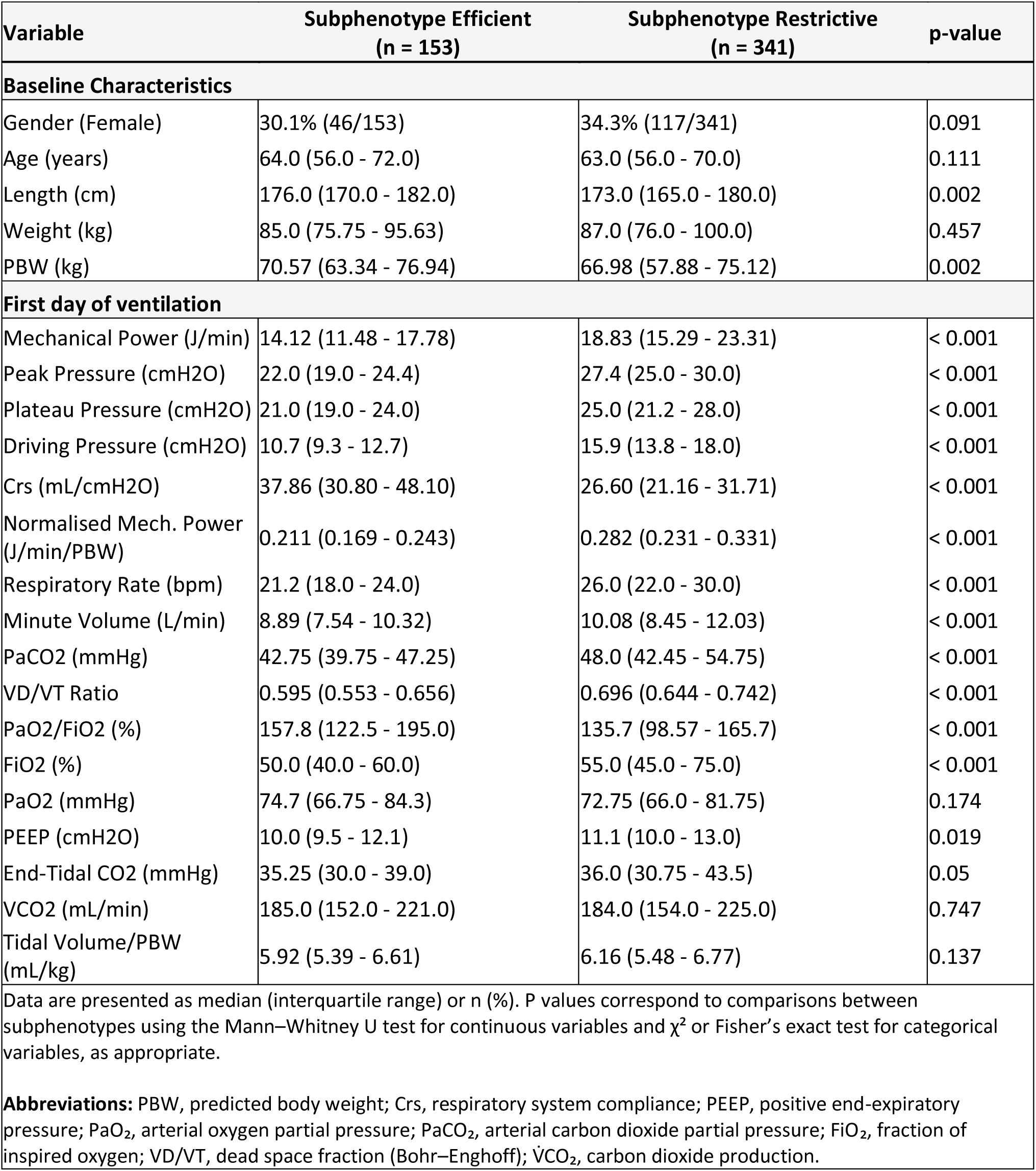
Baseline characteristics and first-day ventilatory and gas-exchange variables by physiological subphenotype in the Amsterdam UMC cohort.

Marked differences were observed in ventilatory mechanics and gas-exchange variables during the first day of invasive mechanical ventilation. Compared with the Efficient subphenotype, patients with the Restrictive subphenotype had higher mechanical power, peak and plateau pressures, DP, RR, minute ventilation (MV), PaCO₂, and dead space fraction, together with lower respiratory system compliance and worse oxygenation (all p < 0.001). Overall, these patterns are consistent with a more mechanically restrictive phenotype with greater ventilatory inefficiency and more severe gas-exchange impairment.

#### 1.8.2 Classification performance of the retrained model

The supervised classifier was retrained in the external validation cohort by replacing VD/VT with ventilatory ratio VR, and using the previously identified GMM–derived labels (Efficient and Restrictive) as reference classes. Principal component analysis demonstrated a clear separation between Efficient and Restrictive patients in the reduced feature space, indicating preservation of the underlying physiological structure after model retraining (Figure 1A). After cross-validated feature selection (using RFECV), the final parsimonious model retained a limited set of physiologically meaningful variables, including DP, RR, VR, nMP, PPlat, PaCO₂, PaO₂/FiO₂, MV, PEEP, and PaO₂ (Figure 1C). SHAP values indicated that DP was the strongest contributor to model predictions, followed by markers of ventilatory demand and efficiency, highlighting the central role of respiratory mechanics and gas-exchange inefficiency in subphenotype discrimination. Model performance was high, with an F1 score of 0.93, sensitivity of 0.92, specificity of 0.97, positive predictive value of 0.93, and negative predictive value of 0.97, indicating balanced and robust classification across physiological subphenotypes (Figure 1B).

**Figure 1.**
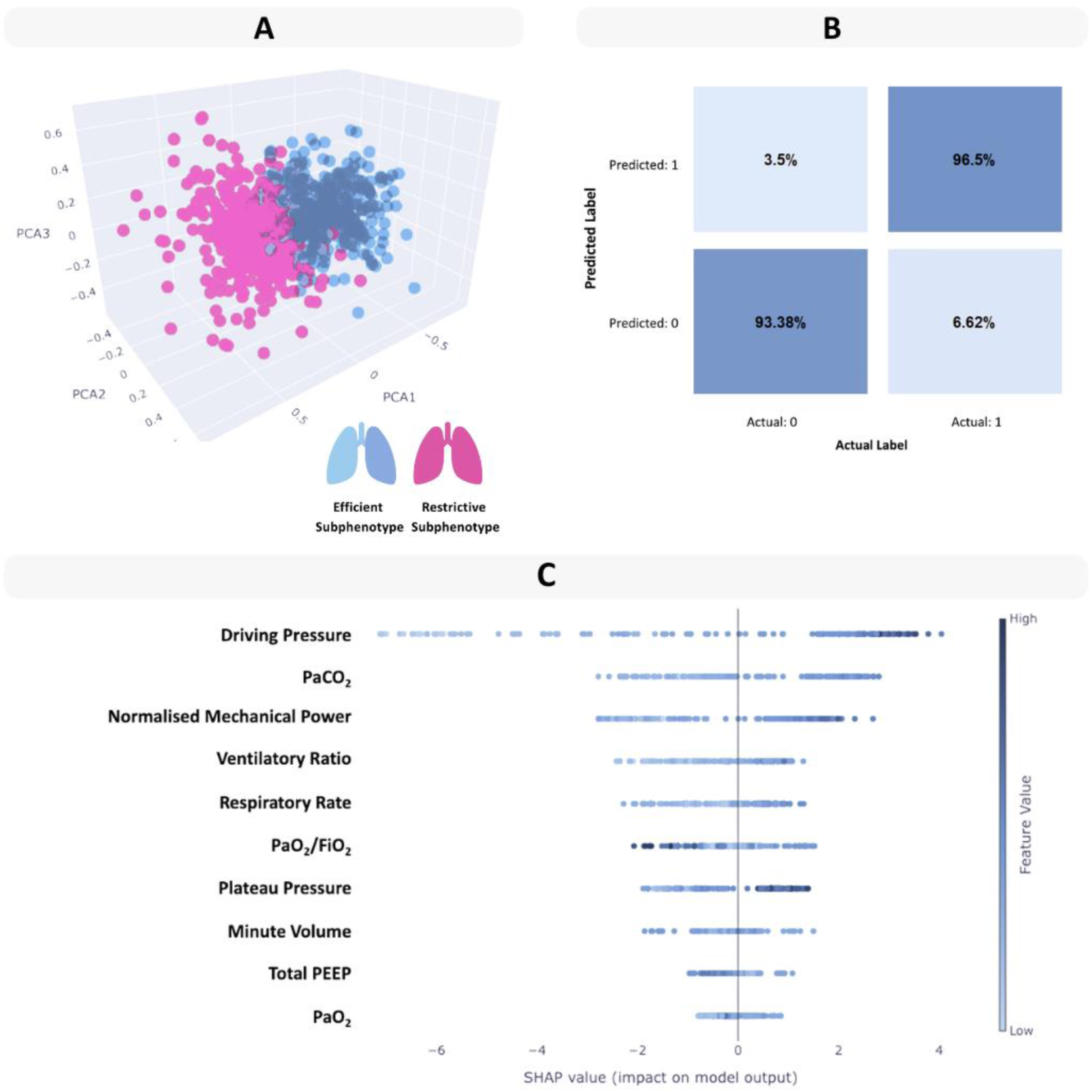
Retraining and performance of the supervised classifier incorporating ventilatory ratio in the Amsterdam UMC cohort. (A) Principal component analysis (PCA) projection demonstrating separation between the Efficient and Restrictive physiological subphenotypes. (B) Row-normalized confusion matrix of the retrained model, summarizing classification performance across subphenotypes. (C) SHAP summary plot showing the relative importance and directional contribution of ventilatory and gas-exchange variables to predictions of the retrained XGBoost model incorporating ventilatory ratio (VR).

### 1.9 Prognostic enrichment across cohorts

Across all cohorts, the Restrictive physiological subphenotype was consistently associated with higher all-cause 28-day mortality compared with the Efficient subphenotype (Figure 2).

**Figure 2.**
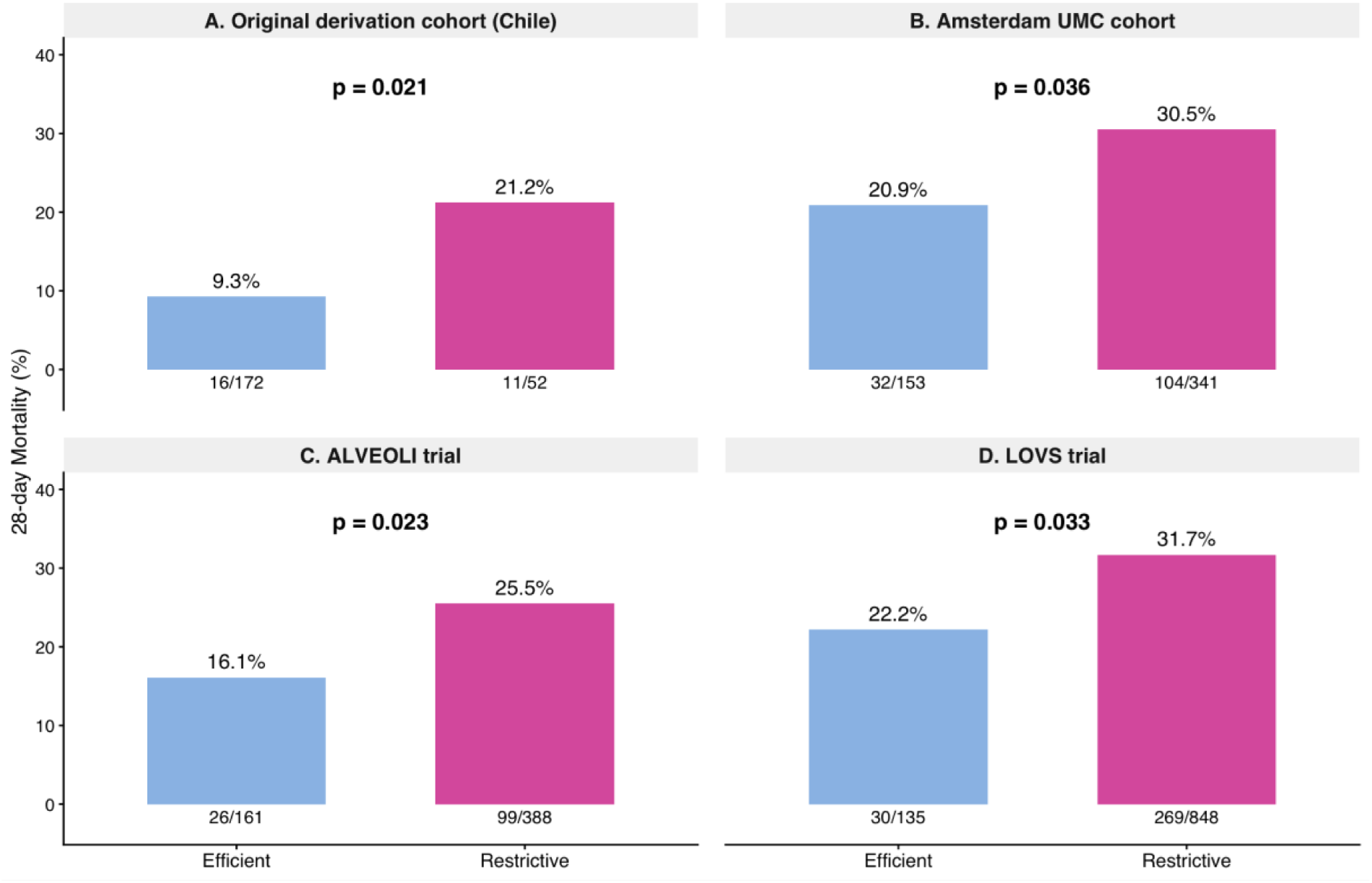
Prognostic enrichment across independent cohorts. Bar plots show 28-day mortality by physiological subphenotype across cohorts: (A) original derivation cohort (Chile), (B) Amsterdam UMC cohort, (C) ALVEOLI trial, and (D) LOVS trial. Percentages above bars indicate 28-day mortality, and numbers below bars indicate deaths/total patients within each subphenotype. P-values correspond to within-cohort comparisons between Efficient and Restrictive subphenotypes.

In the original derivation cohort from Chilean patients, 28-day mortality was significantly higher in Restrictive patients than in Efficient patients (21.2% vs 9.3%, p = 0.021). This prognostic separation was reproduced in the Amsterdam UMC cohort (30.5% vs 20.9%, p = 0.036), as well as in two independent randomised trial populations, including ALVEOLI (25.5% vs 16.1%, p = 0.023) and LOVS (31.7% vs 22.2%, p = 0.033). In ALVEOLI, survival curves further illustrated lower survival in Restrictive patients and a tendency toward poorer survival in Efficient patients exposed to higher PEEP (Supplementary Figure S4).

Baseline ventilatory and gas exchange characteristics differed consistently between physiological subphenotypes in both randomized trial cohorts, with Restrictive patients exhibiting higher DP, RR, and MP, alongside lower oxygenation compared with Efficient patients (Supplementary Tables S1 and S2).

To formally quantify the prognostic impact of physiological subphenotype on mortality, we performed a meta-analysis of study-specific odds ratios comparing the Restrictive versus Efficient subphenotypes across cohorts. In all four cohorts, the Restrictive subphenotype was consistently associated with higher 28-day mortality compared with the Efficient subphenotype.

When pooled across studies, the Restrictive subphenotype was associated with a significantly increased risk of 28-day mortality (pooled OR 1.75, 95% CI 1.36–2.24). There was no evidence of between-study heterogeneity (I² = 0%) (Figure 3), indicating a highly consistent prognostic effect across derivation, validation, and randomized trial cohorts.

**Figure 3.**
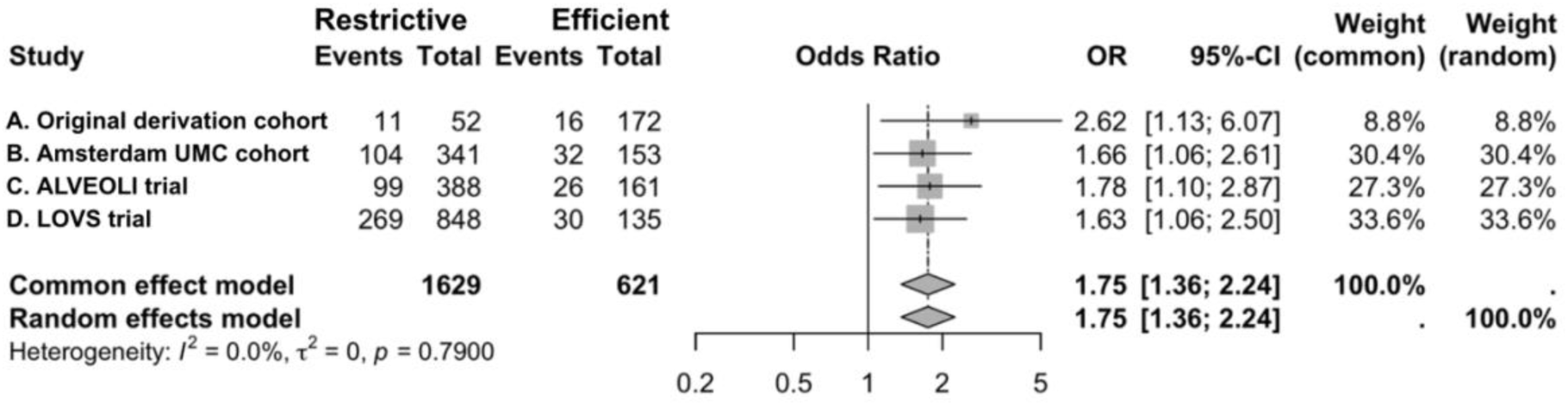
Meta-analysis of 28-day mortality by physiological subphenotype. Forest plot showing study-specific and pooled odds ratios for 28-day mortality comparing Restrictive versus Efficient physiological subphenotypes across cohorts. A meta-analysis demonstrated a consistently higher mortality risk in the Restrictive subphenotype (pooled OR 1.75, 95% CI 1.36–2.24), with no evidence of between-study heterogeneity (I² = 0%).

**Figure 4.**
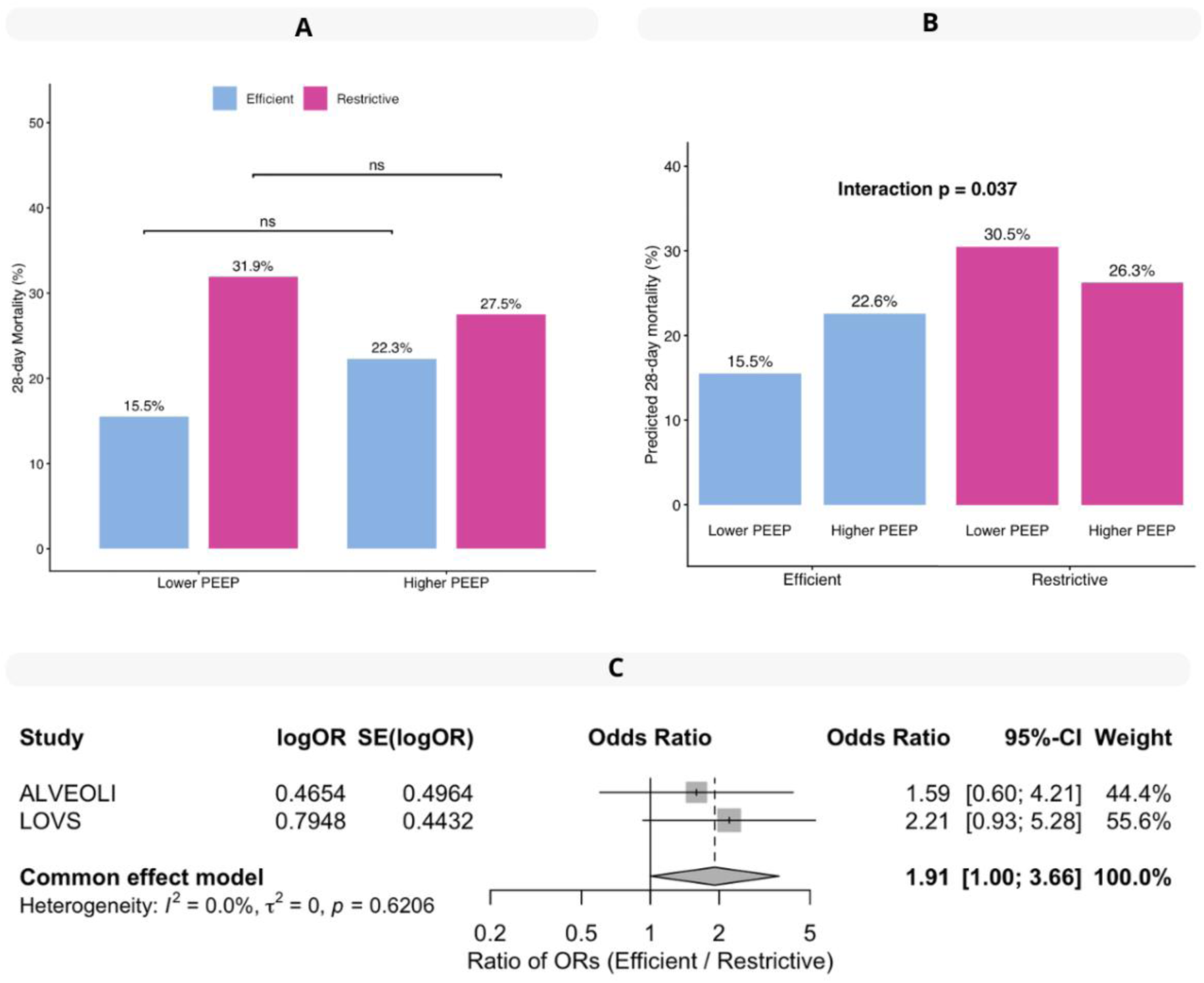
Predictive enrichment by physiological subphenotypes. **A.** Observed 28-day mortality by PEEP strategy stratified by physiological subphenotype. Bar plots show crude 28-day mortality under lower and higher PEEP strategies in the combined ALVEOLI and LOVS cohorts, stratified by Efficient and Restrictive physiological subphenotypes. Percentages above bars indicate 28-day mortality, and numbers below bars indicate deaths/total patients within each subphenotype. P-values correspond to within-subphenotype comparisons between PEEP strategies. **B.** Adjusted 28-day mortality by physiological subphenotype and PEEP strategy. Bar plots show predicted 28-day mortality probabilities derived from an adjusted logistic regression model fitted in the combined ALVEOLI and LOVS cohorts. Estimates are shown for lower and higher PEEP within each physiological subphenotype. The interaction p-value reflects statistical evidence of effect modification between PEEP strategy and subphenotype. **C.** Differential effect of higher PEEP according to physiological subphenotype (interaction analysis) Forest plot displays the pooled interaction effect comparing the odds ratios of higher versus lower PEEP between Efficient and Restrictive physiological subphenotypes, estimated using a fixed-effects two-stage individual patient data meta-analysis. The vertical dashed line indicates an odds ratio of 1.

### 1.10 Predictive enrichment across cohorts

Predictive enrichment for PEEP strategy was evaluated using individual patient data from the ALVEOLI and LOVS randomised trials, comprising a total of 1,532 patients (ALVEOLI, n = 549; LOVS, n = 983), all classified into physiological subphenotypes using the supervised model prior to randomisation. Randomization to lower PEEP or higher PEEP strategies according to the original trial was evaluated in an intention to treat analysis.

Evidence of heterogeneity of treatment effect by physiological subphenotype was observed in the one-stage IPD analysis (Figure 3B; Supplementary Table S6). A statistically significant interaction between PEEP strategy and subphenotype was observed (p for interaction = 0.037), indicating that the effect of higher versus lower PEEP differed according to physiological subphenotype. Consistent results were obtained in a fixed-effects two-stage IPD meta-analysis, which also showed statistical evidence for heterogeneity of treatment effect between subphenotypes (interaction OR 1.91, 95% CI 1.00–3.66; p = 0.04; Figure 3C), with no between-study heterogeneity (I² = 0%).

The direction of the treatment effect was consistent across analytical approaches. In crude pooled analyses, higher PEEP was associated with higher mortality among Efficient patients and lower mortality among Restrictive patients, although these differences did not reach statistical significance (Figure 3A; Supplementary Tables S4 and S5). These opposing directions were preserved in the adjusted one-stage IPD model and were consistently observed in trial-specific and pooled estimates from the two-stage IPD meta-analysis (Supplementary Figures S7 and S8). In both ALVEOLI and LOVS, higher PEEP showed concordant trends towards harm in Efficient patients and relative benefit in Restrictive patients, supporting the robustness and biological coherence of the observed effect modification.

## Discussion

We confirmed the clinical relevance of classifying ARDS into Efficient and Restrictive physiological subphenotypes across three independent validation cohorts. Key ventilatory and gas-exchange variables were consistently and significantly different between subphenotypes, reproducing the physiological patterns originally identified in the derivation cohort (13). Importantly, these differences were observed despite heterogeneity in geography and clinical practice, supporting the robustness and generalisability of the physiological phenotyping approach. Across all studied populations, mortality rates were consistently higher in patients classified as Restrictive compared with those classified as Efficient, confirming the prognostic relevance of the physiological subphenotypes. Furthermore, analyses of individual patient data from two landmark RCTs suggested HTE for PEEP strategy between physiological subphenotypes. In both trials, the direction of treatment effects was concordant, with higher PEEP tending to be associated with improved outcomes in Restrictive patients and worse outcomes in Efficient patients. These findings were supported by both one-stage and two-stage IPD analyses, including a pooled interaction estimate indicating statistically significant effect modification.

### 1.11 External validation and physiological interpretation of ARDS subphenotypes

Across cohorts, we replicated the key finding from the derivation cohort; Efficient and Restrictive physiological subphenotypes represent two distinct patterns of lung mechanics and gas exchange behaviour in ARDS. Restrictive patients consistently exhibited a more mechanically severe profile, characterised by higher driving pressures, reduced respiratory system compliance, increased ventilatory demand, and impaired ventilatory efficiency, consistent with substantial ventilation–perfusion mismatch reflected by worse oxygenation and increased physiological dead space (26,27). Together, these routinely available bedside measurements reflect underlying pathophysiological mechanisms (4) compatible with more diffuse alveolar involvement, increased shunt fraction, reduced functional lung size (“baby lung”), and higher mechanical stress for a given ventilatory load (14,28). By contrast, the Efficient subphenotype was characterised by relatively preserved respiratory mechanics and gas exchange, suggesting a larger functional lung size and lower physiological impairment despite meeting ARDS diagnostic criteria and inclusion in major interventional studies(1,29).

This physiological framework overlaps conceptually with previously described ARDS subphenotypes based on recruitability, morphology, and biological response, while offering important advantages in feasibility and clinical applicability. Recruitable versus non-recruitable subphenotypes identified using respiratory mechanics, gas exchange, and computed tomography showed similar patterns of reduced compliance and worse oxygenation in patients with diffuse lung involvement (30,31), but relied on complex imaging protocols and recruitment manoeuvres that limit scalability and external validation. Biological subphenotypes, such as the hypo- and hyperinflammatory subtypes described by Calfee and colleagues, demonstrated important biological heterogeneity (32) and may partially overlap with the physiological subphenotypes, with hypoinflammatory features aligning more closely with Efficient patients and hyperinflammatory features with Restrictive patients, as suggested by differences in clinical characteristics and outcomes (9). However, a key limitation of this approach is that circulating plasma biomarkers primarily reflect systemic inflammation and do not necessarily capture lung-specific pathophysiology (33), and similar inflammatory subphenotypes have been identified in patients without ARDS (34). Radiological classifications have likewise identified non-focal ARDS as a mechanically more severe subphenotype, with studies showing that non-focal morphology is associated with higher respiratory system elastance and worse oxygenation, findings comparable to the Restrictive subphenotype identified in the present study (35). However, the clinical utility of radiological classification has been limited by substantial interobserver variability and misclassification, which may partly explain the lack of benefit observed in personalised ventilation strategies in intention-to-treat analyses (20). In contrast, the present approach enables reproducible, operator-independent physiological subphenotyping across multiple cohorts using simple bedside variables, in line with ESICM recommendations (10), and provides a robust foundation for examining prognostic differences and differential responses to ventilatory interventions, which we explored through analyses of 28-day mortality and HTE according to PEEP strategy.

### 1.12 Prognostic relevance of physiological ARDS subphenotypes

Across all cohorts, classification into Efficient and Restrictive physiological subphenotypes was consistently associated with clinically meaningful differences in outcomes, with higher all-cause 28-day mortality observed among patients classified as Restrictive. These differences were not driven by a single variable, but rather reflected the integrated physiological profile defining each subphenotype.

The poorer prognosis of the Restrictive subphenotype is physiologically plausible and consistent with established determinants of outcome in ARDS. Key features of this subphenotype, including higher driving pressure, reduced respiratory system compliance, impaired oxygenation, increased ventilatory demand, and elevated physiological dead space, have each been independently associated with increased mortality (11,26,27,36). By integrating these dimensions into a unified physiological profile, subphenotyping provides a more comprehensive representation of disease severity than traditional prognostic stratification based on oxygenation severity or single mechanical ventilation variables. Although indices such as the PaO₂/FiO₂ ratio are widely used for ARDS classification and risk stratification (1,26), baseline oxygenation alone incompletely reflects lung mechanics, ventilatory efficiency, and functional lung size, and has shown limited prognostic performance in acute hypoxemic respiratory failure (37,38). In contrast, physiological subphenotyping offers clinically interpretable prognostic enrichment grounded in pathophysiology and raises the possibility that baseline physiological profiles may also modify responses to ventilatory strategies, which we explored through analyses of HTE related to PEEP strategy.

### 1.13 Predictive enrichment and heterogeneity of treatment effect

A statistically significant interaction between physiological subphenotype and PEEP strategy was observed for all-cause 28-day mortality, indicating heterogeneity of treatment effect according to baseline physiological profile. This interaction was consistently identified across analytical approaches, including study-adjusted analyses and IPD meta-analyses using both one-stage and two-stage frameworks, supporting the presence of differential treatment effects rather than random variation.

Given the presence of a significant interaction, subgroup analyses suggested divergent responses to PEEP strategy between subphenotypes. Patients classified as Restrictive tended to have improved outcomes when managed with higher PEEP, whereas patients classified as Efficient showed a tendency toward worse outcomes under higher PEEP exposure. These opposing directions are physiologically coherent with the defining characteristics of each subphenotype. Restrictive patients, characterised by impaired respiratory system compliance and increased shunt and physiological dead space, exhibit a physiological profile consistent with reduced functional lung size and greater recruitability, and may therefore benefit from higher PEEP through alveolar recruitment and stabilisation of dependent lung units (39,40). In contrast, Efficient patients, with relatively preserved respiratory mechanics and lower indices of ventilatory inefficiency, may be more susceptible to overdistension and increased mechanical stress when exposed to higher PEEP levels, particularly in the absence of substantial recruitable lung (41,42).

Previous attempts to identify heterogeneity of treatment effect in ARDS have yielded inconsistent results. Biological subphenotypes based on hypo- and hyperinflammatory profiles initially suggested differential responses to PEEP (32,43), however, these findings have not been consistently replicated across independent cohorts (19). Similarly, stratification based on oxygenation using the PaO₂/FiO₂ ratio may capture differences in baseline severity (26), but has not been formally established as a predictive enrichment strategy for PEEP responsiveness (44), nor does it account for underlying lung mechanics or recruitability. Recruitment-based subphenotypes derived from imaging or extended recruitment manoeuvres conceptually align with differential PEEP response (30,39), but their clinical applicability is limited by the requirement for advanced imaging, protocolised manoeuvres, and specialised expertise, which are frequently unavailable in routine practice. Across these different approaches, a common limitation is the lack of an integrated, bedside representation of respiratory physiology. Evidence from longitudinal respiratory phenotyping in acute respiratory failure due to COVID-19 has also described restrictive physiological trajectories associated with distinct ventilatory patterns, reinforcing the broader concept that baseline respiratory physiology may modify treatment response (45).

In this context, physiological subphenotyping based on routinely available ventilatory and gas-exchange variables offers a pragmatic alternative for predictive enrichment. By capturing integrated properties of respiratory mechanics and ventilatory efficiency, this approach provides a physiologically interpretable basis for examining differential responses to ventilatory interventions.

### 1.14 Limitations and future directions

The findings of this study should be interpreted in the context of several considerations. Evidence for HTE was derived from post hoc analyses of existing randomized trials. Although the interaction between physiological subphenotype and PEEP strategy was consistent across analytical approaches, the observed effect size was modest.

Baseline ventilatory settings may have influenced subphenotype classification. As PEEP was included as a model variable and also directly affects key physiological parameters such as compliance, oxygenation, and ventilatory efficiency, the identified subphenotypes likely reflect both intrinsic pulmonary physiology and clinician-driven optimisation of ventilatory support. This should be considered when interpreting both prognostic associations and observed HTE. Nonetheless, by integrating multiple complementary physiological variables, this approach provides a robust and physiologically representative characterisation of respiratory dysfunction.

Importantly, randomized trials differ in how high and low PEEP strategies are defined, and patients with ARDS exhibit substantial underlying physiological heterogeneity. Together, these factors increase the difficulty of detecting treatment effects in unselected ARDS populations. Physiological subphenotyping represents one possible approach to improve predictive enrichment, but requires further validation. While physiological subphenotypes were externally validated across independent observational and trial cohorts, HTE was identified in the ALVEOLI and LOVS trials; however, the magnitude and precision of the treatment effect remain limited, with estimates close to the threshold of statistical significance. Further validation of these findings in independent randomised datasets is therefore essential.

Importantly, such validation should be guided by the consistency assumption, a key requirement for causal interpretation in HTE analyses (46), which states that treatment effects can only be meaningfully compared when interventions and control conditions are sufficiently similar across studies.

Based on this principle, we hypothesise that the EXPRESS trial (47) represents the most appropriate dataset for further evaluation. Several key differences in trial design support this choice. First, the lower PEEP arm in EXPRESS resulted in a PEEP most comparable to LOVS and ALVEOLI (16), despite the use of a different protocol for setting PEEP. The lower PEEP arm in the other studies potentially available for external validation (ART, PHARLAP and OLA) (48–50) also used the lower PEEP-FiO2 table in the control arm, but this resulted in a higher PEEP which could not be completely explained by the baseline PaO2/FiO2.

Implemention of the PEEP-FiO2 table was likely different across trials, with very specific and protocolised adjustments outlined in ALVEOLI and much less strict guidance in the other studies (either no guidance available, or with different steps outlined in the protocol), which may further explain variation in actual PEEP settings. Second, in the experimental arms of the ART, PHARLAP, and OLA trials, a prolonged RM was used. We assume that the HTE across subphenotypes hypothesized in our study will not transport to trials that relied on these prolonged RMs as this may have harmful effects independent of the subphenotypes described in this study and would therefore reduce the hypothesized HTE. These differences may influence the effective physiological exposure to PEEP and reduce comparability across studies, thereby challenging the consistency assumption.

Pre-specifying the expected presence or absence of HTE across datasets is essential to avoid post hoc interpretation. Although HTE was identified using both one-stage and two-stage IPD approaches, replication in an independent cohort is required to confirm the robustness of the effect, particularly given the borderline magnitude of the observed interaction.

## Conclusion

Efficient and Restrictive subphenotypes were consistently replicated across cohorts, and were consistently associated with distinct patterns of respiratory mechanics, gas exchange, and clinical outcomes across independent cohorts, supporting their physiological and prognostic relevance. We identified HTE for PEEP strategy, with a significant interaction between subphenotype and the clinical response to a low or high PEEP strategy. Taken together, these results support physiological subphenotyping as a practical and clinically interpretable framework to address heterogeneity in ARDS. Further validation in additional randomized trials is required to determine whether this approach can be used for predictive enrichment and to guide personalised ventilatory strategies in clinical practice.

## Data Availability

All data produced in the present study are available upon reasonable request to the authors

