## Supplementary Material for "Physiological subphenotypes of ARDS: Prognostic and predictive enrichment for PEEP strategy"

Supplementary Figure S1. Overview of model development, external validation, and application to treatment response

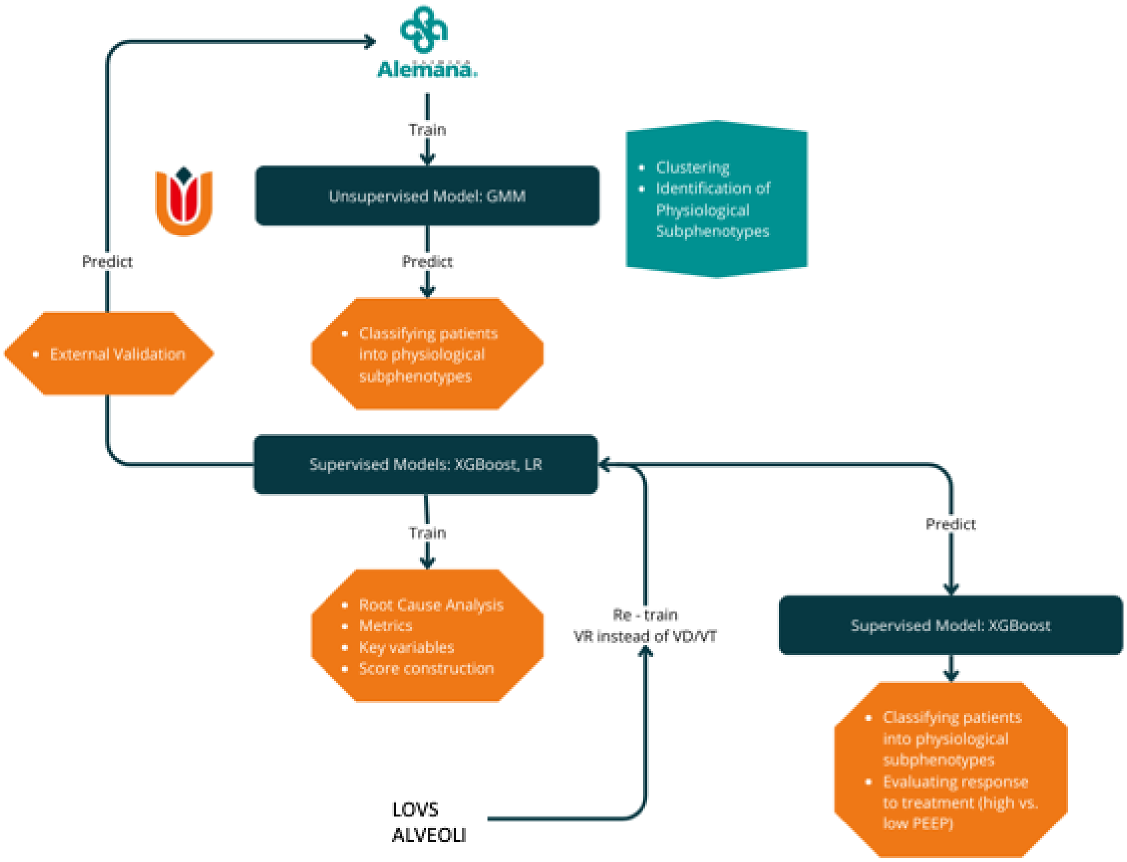


Schematic overview of the analytical workflow. An unsupervised Gaussian Mixture Model was used to identify physiological subphenotypes based on ventilatory and gas-exchange variables. These labels were then used to train supervised classifiers (XGBoost and logistic regression) for parsimonious bedside application. The resulting models were externally validated and subsequently applied to randomized trial datasets to assess clinical outcomes and heterogeneity of treatment effect according to PEEP strategy.

Supplementary Figure S2. Selection of the optimal number of features for the parsimonious XGBoost classifier

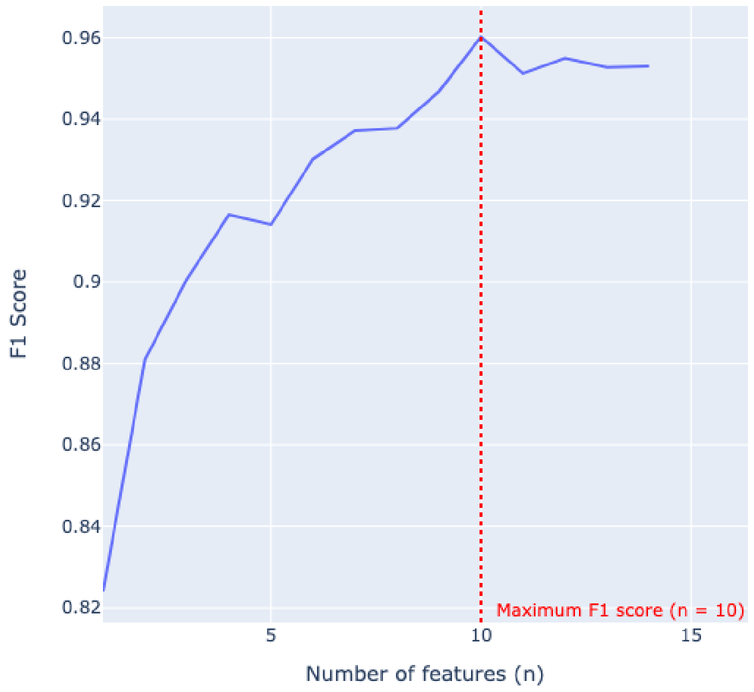


XGBoost classifier performance (F1 score) across increasing numbers of selected features. The dashed line indicates the parsimonious model achieving maximal performance with ten features.

Supplementary Figure S3. Feature importance of the XGBoost classifier using ventilatory ratio

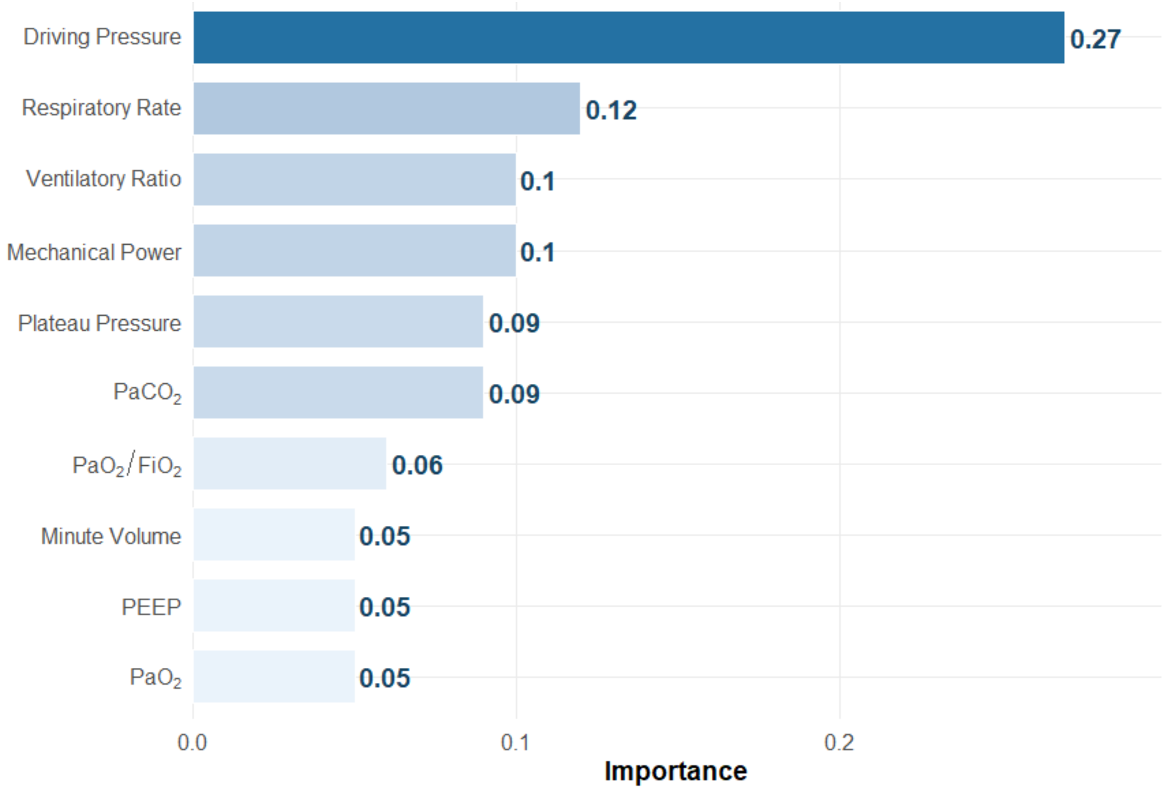


Relative importance of ventilatory and gas-exchange variables in the XGBoost classifier trained to reproduce GMM-derived physiological subphenotypes. Driving pressure and respiratory rate were the most influential predictors.

Supplementary Figure S4. Survival by physiological subphenotype and PEEP strategy in the ALVEOLI trial

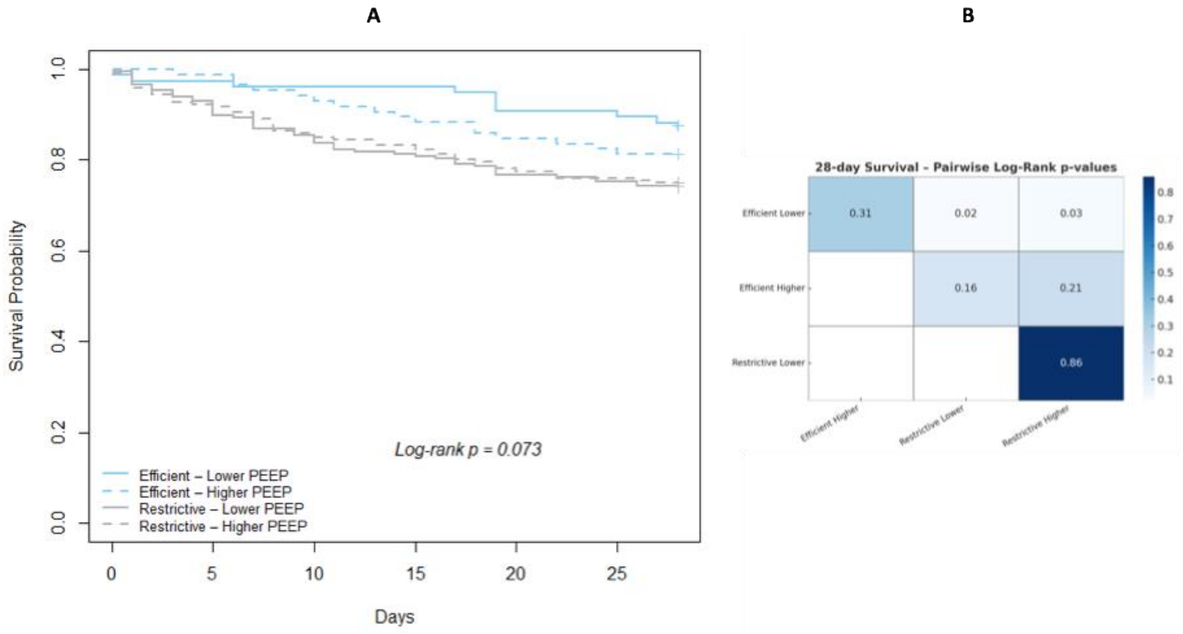


(A) Kaplan–Meier survival curves stratified by physiological subphenotype (Efficient vs Restrictive) and PEEP strategy (higher vs lower PEEP) in the ALVEOLI trial.
(B) Pairwise log-rank p-values for 28-day survival comparing subphenotype–PEEP groups.
Survival differed across groups, with separation primarily driven by physiological subphenotype, while survival was similar between Restrictive patients managed with higher versus lower PEEP.

Supplementary Figure S5. Ventilator-free days by physiological subphenotype and PEEP strategy in the ALVEOLI trial.

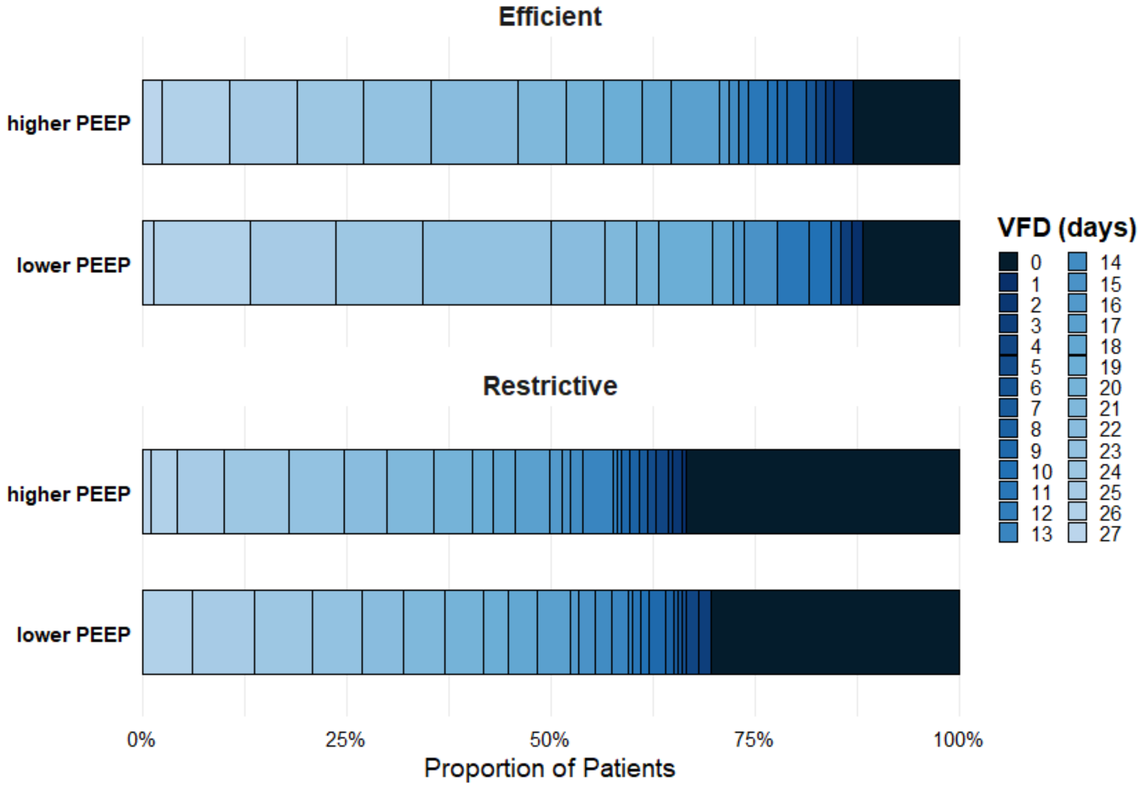


Distribution of ventilator-free days at 28 days stratified by physiological subphenotype (Efficient vs Restrictive) and PEEP strategy (higher vs lower PEEP).

Supplementary Figure S6. In-hospital mortality by physiological subphenotype in the ALVEOLI trial.

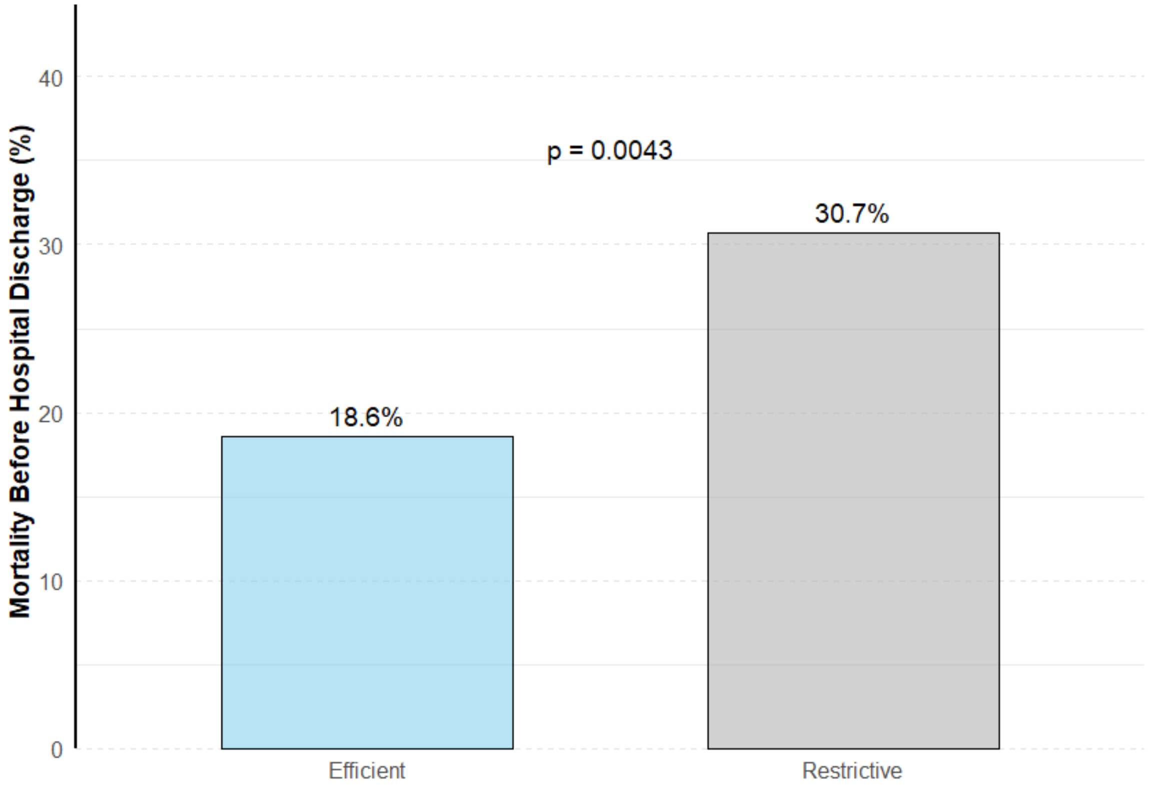


In-hospital mortality rates stratified by physiological subphenotype.

Supplementary Figure S7. Effect of higher versus lower PEEP on 28-day mortality in the Efficient physiological subphenotype

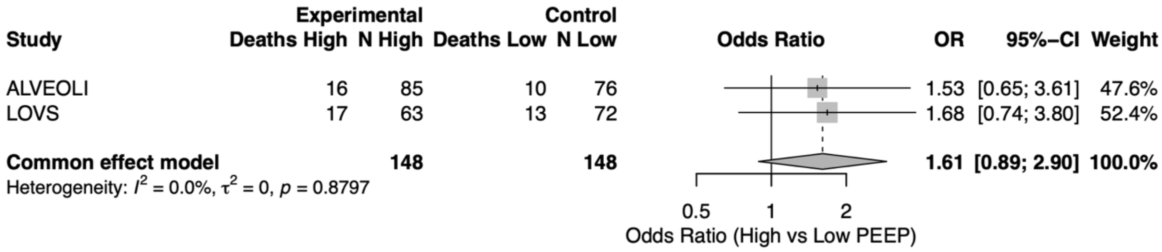


Forest plot showing study-specific and pooled odds ratios for 28-day mortality comparing higher versus lower PEEP in patients classified as the Efficient physiological subphenotype across the ALVEOLI and LOVS trials. Odds ratios were pooled using a fixed-effects individual patient data meta-analysis.

Supplementary Figure S8. Effect of higher versus lower PEEP on 28-day mortality in the Restrictive physiological subphenotype

| Study | Experimental Deaths High | Experimental N High | Control Deaths Low | Control N Low | OR | 95%-CI | Weight |
|---|---|---|---|---|---|---|---|
| ALVEOLI | 48 | 191 | 51 | 197 | 0.96 | [0.61; 1.52] | 28.8% |
| LOVS | 118 | 412 | 151 | 436 | 0.76 | [0.57; 1.01] | 71.2% |
| **Common effect model** | | **603** | | **633** | **0.81** | **[0.63; 1.04]** | **100.0%** |

Heterogeneity: $I^2 = 0.0\%$, $\tau^2 = 0$, $p = 0.3892$

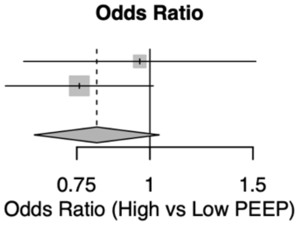


Forest plot showing study-specific and pooled odds ratios for 28-day mortality comparing higher versus lower PEEP in patients classified as the Restrictive physiological subphenotype across the ALVEOLI and LOVS trials. Odds ratios were pooled using a fixed-effects individual patient data meta-analysis.

Supplementary Table S1. XGBoost hyperparameters

| Parameter | Value |
|---|---|
| n_estimators | 100 |
| max_depth | 4 |
| learning_rate | 0.1 |
| subsample | 0.8 |
| colsample_bytree | 0.8 |
| reg_alpha | 0.01 |
| reg_lambda | 1 |
| objective | binary:logistic |
| random_state | 55 |

Hyperparameters used for training the XGBoost classifier, including model architecture, regularisation, and optimisation settings.

Supplementary Table S2. Baseline ventilatory and gas-exchange characteristics by physiological subphenotype in the ALVEOLI trial

| Variable | Efficient (N = 161) | Restrictive (N = 388) | p-value |
|---|---|---|---|
| Driving Pressure ($cmH_2O$) | 12.00 (10.00 – 13.00) | 19.00 (16.00 – 22.00) | <0.001 |
| Minute Ventilation (L/min) | 8.34 (7.00 – 10.00) | 10.00 (8.32 – 12.80) | <0.001 |
| Ventilatory Ratio | 0.82 (0.63 – 0.97) | 1.04 (0.82 – 1.36) | <0.001 |
| $PaO_2$ / $FiO_2$ Ratio | 162.22 (115.00 – 210.00) | 122.54 (92.00 – 166.00) | <0.001 |
| Mechanical Power / PBW (J/min/kg) | 0.24 (0.18 – 0.32) | 0.42 (0.30 – 0.56) | <0.001 |
| Plateau Pressure ($cmH_2O$) | 20.00 (18.00 – 22.00) | 30.00 (25.00 – 32.00) | <0.001 |
| PEEP ($cmH_2O$) | 8.00 (5.00 – 10.00) | 10.00 (8.00 – 12.00) | <0.001 |
| Respiratory Rate (bpm) | 16.00 (14.00 – 20.00) | 20.00 (16.00 – 28.00) | <0.001 |
| $PaCO_2$ (mmHg) | 37.00 (32.00 – 40.00) | 39.00 (33.50 – 44.00) | 0.005 |
| $PaO_2$ (mmHg) | 80.00 (68.00 – 99.00) | 75.00 (66.00 – 90.00) | 0.021 |

Values are presented as median (interquartile range). Comparisons between Efficient and Restrictive physiological subphenotypes were performed using the Wilcoxon rank-sum test. PBW indicates predicted body weight.

Supplementary Table S3. Baseline ventilatory and gas-exchange characteristics by physiological subphenotype in the LOVS trial

| Variable | Efficient (N = 135) | Restrictive (N = 848) | p-value |
|---|---|---|---|
| Driving Pressure ($cmH_2O$) | 11.00 (9.50 – 12.00) | 19.00 (16.00 – 22.00) | <0.001 |
| Minute Ventilation (L/min) | 9.60 (7.65 – 11.65) | 11.30 (9.30 – 13.89) | <0.001 |
| Ventilatory Ratio | 0.94 (0.91 – 0.96) | 0.94 (0.87 – 0.95) | 0.621 |
| $PaO_2$ / $FiO_2$ Ratio | 170.00 (133.50 – 210.00) | 154.50 (110.00 – 185.75) | 0.001 |
| Mechanical Power / PBW (J/min/kg) | 0.23 (0.18 – 0.29) | 0.36 (0.28 – 0.46) | <0.001 |
| Plateau Pressure ($cmH_2O$) | 22.00 (19.00 – 24.00) | 30.00 (28.00 – 34.00) | <0.001 |
| PEEP ($cmH_2O$) | 10.00 (10.00 – 12.50) | 10.00 (10.00 – 14.00) | 0.215 |
| Respiratory Rate (bpm) | 18.00 (15.00 – 22.00) | 22.00 (18.00 – 27.00) | <0.001 |
| $PaCO_2$ (mmHg) | 39.00 (35.25 – 45.00) | 41.00 (36.00 – 48.00) | 0.025 |
| $PaO_2$ (mmHg) | 79.00 (68.00 – 88.00) | 76.00 (65.00 – 90.00) | 0.428 |

Values are presented as median (interquartile range). Comparisons between Efficient and Restrictive physiological subphenotypes were performed using the Wilcoxon rank-sum test. PBW indicates predicted body weight.

Supplementary Table S4. Descriptive 28-day mortality by physiological subphenotype and PEEP strategy

| Dataset | Subphenotype | High PEEP | Low PEEP |
|---|---|---|---|
| ALVEOLI | Efficient | 16/85 (18.8%) | 10/76 (13.2%) |
| LOVS | Efficient | 17/63 (27.0%) | 13/72 (18.1%) |
| ALVEOLI | Restrictive | 48/191 (25.1%) | 51/197 (25.9%) |
| LOVS | Restrictive | 118/412 (28.6%) | 151/436 (34.6%) |

Values are shown as number of deaths / total patients (%).

Supplementary Table S5. Descriptive 28-day mortality by physiological subphenotype and PEEP strategy (all trials combined)

| Subphenotype | High PEEP | Low PEEP |
|---|---|---|
| **Efficient** | 33 / 148 (22.3%) | 23 / 148 (15.5%) |
| **Restrictive** | 166 / 603 (27.5%) | 202 / 633 (31.9%) |

Values are shown as number of deaths / total patients (%).

Supplementary Table S6. One-stage IPD logistic regression model for 28-day mortality

Outcome: All-cause 28-day mortality
Population: ALVEOLI and LOVS trials (n = 1,532)
Model: Logistic regression (binomial, logit link)
Adjustment: Trial (dataset)

| Variable | β | SE | OR | 95% CI | p-value |
|---|---|---|---|---|---|
| Intercept | −1.860 | 0.237 | 0.16 | 0.10–0.24 | <0.001 |
| Restrictive vs Efficient subphenotype | 0.874 | 0.244 | 2.40 | 1.51–3.95 | <0.001 |
| Higher vs Lower PEEP strategy | 0.466 | 0.302 | 1.59 | 0.89–2.91 | 0.33 |
| LOVS vs ALVEOLI trial | 0.326 | 0.126 | 1.38 | 1.08–1.77 | 0.010 |
| **Higher PEEP × Restrictive subphenotype (interaction)** | **−0.675** | **0.327** | **0.51** | **0.27–0.96** | **0.037** |

The model includes treatment group (higher vs lower PEEP), physiological subphenotype, their interaction term, and adjustment for trial membership. Odds ratios are reported with 95% confidence intervals. Statistical significance of the interaction was assessed using likelihood ratio tests.

Supplementary Table S7. Study-specific and pooled odds ratios for higher versus lower PEEP stratified by physiological subphenotype.

| Subphenotype | Study | OR (High vs Low PEEP) | 95% CI | p-value |
|---|---|---|---|---|
| Efficient | ALVEOLI | 1.53 | 0.65–3.61 | 0.33 |
| | LOVS | 1.68 | 0.74–3.80 | 0.22 |
| | Pooled | 1.61 | 0.89–2.90 | 0.11 |
| Restrictive | ALVEOLI | 0.96 | 0.61–1.52 | 0.86 |
| | LOVS | 0.76 | 0.57–1.01 | 0.06 |
| | Pooled | 0.81 | 0.63–1.04 | 0.09 |
| **Interaction (Eff – Restr)** | **Pooled** | **1.91** | **1.00–3.66** | **0.04** |

Odds ratios were estimated using study-specific logistic regression models and pooled using a fixed-effects two-stage individual patient data meta-analysis. The interaction estimate represents the ratio of odds ratios comparing the effect of higher versus lower PEEP between Efficient and Restrictive subphenotypes.
